# COVID-19 incidence and mortality in the Metropolitan Region, Chile: time, space, and structural factors

**DOI:** 10.1101/2020.09.15.20194951

**Authors:** Pablo Villalobos Dintrans, Claudio Castillo, Felipe De La Fuente, Matilde Maddaleno

## Abstract

Chile has been heavily affected by the COVID-19 pandemic. This article analyzes the association of different groups of factors—demographic, health-related, and socioeconomic—on COVID-19-related outcomes. Using the municipalities of the Metropolitan Region the study looks at the role of time dynamics, space and place in cases and deaths during a 100-days period.

Results show that common and idiosyncratic elements that explain the prevalence and dynamics of infections and mortality, with an important role of social determinants of health, particularly multidimensional poverty index and use of public transportation, in explaining differences in outcomes.

The article contributes to the understanding of the determinants of COVID-19 outcomes in a specific region, but also highligths the need to consider time-space dynamics and social determinants as key in the analysis. The results are specially relevant for similar research in unequal settings.

## Introduction

The novel coronavirus, known as Severe Acute Respiratory Syndrome 2 (SARS-Cov2) firstly described in China at the end of 2019, has produced the new coronavirus disease (COVID-19), declared as a pandemic by the World Health Organization on January 30 2020 (WHO) (1,2). By 31 July 2020, this pandemic has caused over seventeen million confirmed cases and 668.910 deaths worldwide (3).

The region of the Americas is currently the center of the pandemic and Chile has been one of the countries more affected by this new virus (3). Despite the efforts in terms of testing and the application of several measures to stop infection (4,5), the number of cases reached 395.261 by 30 July 2020, and the country has one of the highest mortality per population in the world (6,7). In this context, understanding the factors behind the spread of the virus and the high mortality rates in the country are relevant, particularly considering that the COVID-19 is an ongoing challenge for policymakers.

Several studies on the determinants of COVID-19 have highlighted the multicausal nature of the disease. Research has investigated the impact of countries’ demographic profile, the effect of preexisting health conditions, and the impact of socioeconomic factors in explaining COVID-19-related outcomes (8-13). This is also the case for Chile, where reports have shown how demographics and health conditions relate to COVID-19 outcomes (13). However, the links with social determinants of health have been less studied in the country. Moreover, other relevant features of the disease, such as its temporal and spatial dynamics, are almost completely absent.

This study aims to analyze the association of different groups of factors—demographic, health-related, and socioeconomic—on COVID-19 outcomes: infections and deaths. For the analysis, we use the 52 administrative units (municipalities) in the Metropolitan Region (MR), Chile.

The focus on the MR area is justified by several reasons. First, in terms of scale, this is a highly-populated area (the largest in Chile), which includes the country’s capital, Santiago, and where 40.5% of the Chilean population lives (14). Second, in terms of the impact of the COVID-19, by July 31 2020, the MR clusters most of the cases in the country with 71.91% of total cases and 74.58% of national deaths (13); compared to the national-level reality, the region also exhibits larger cumulative incidence rates (3491.3 vs 2031.3) and mortality rates per 100,000 people (63.7 vs. 30.7) (14). Both features make this region particularly interesting in terms of research and health-care policies. Finally, the Metropolitan area has particular spatial characteristics that makes it an interesting case of analysis. On the one hand, the 52 municipalities are spread over a relatively small area, with most municipalities highly connected, creating an area where administrative boundaries exist but within a large conurbation area. As shown in Fig 1, several municipalities share a small area around the center of the region, surrounding downtown Santiago (the clearer area in the middle).

**Fig 1.**
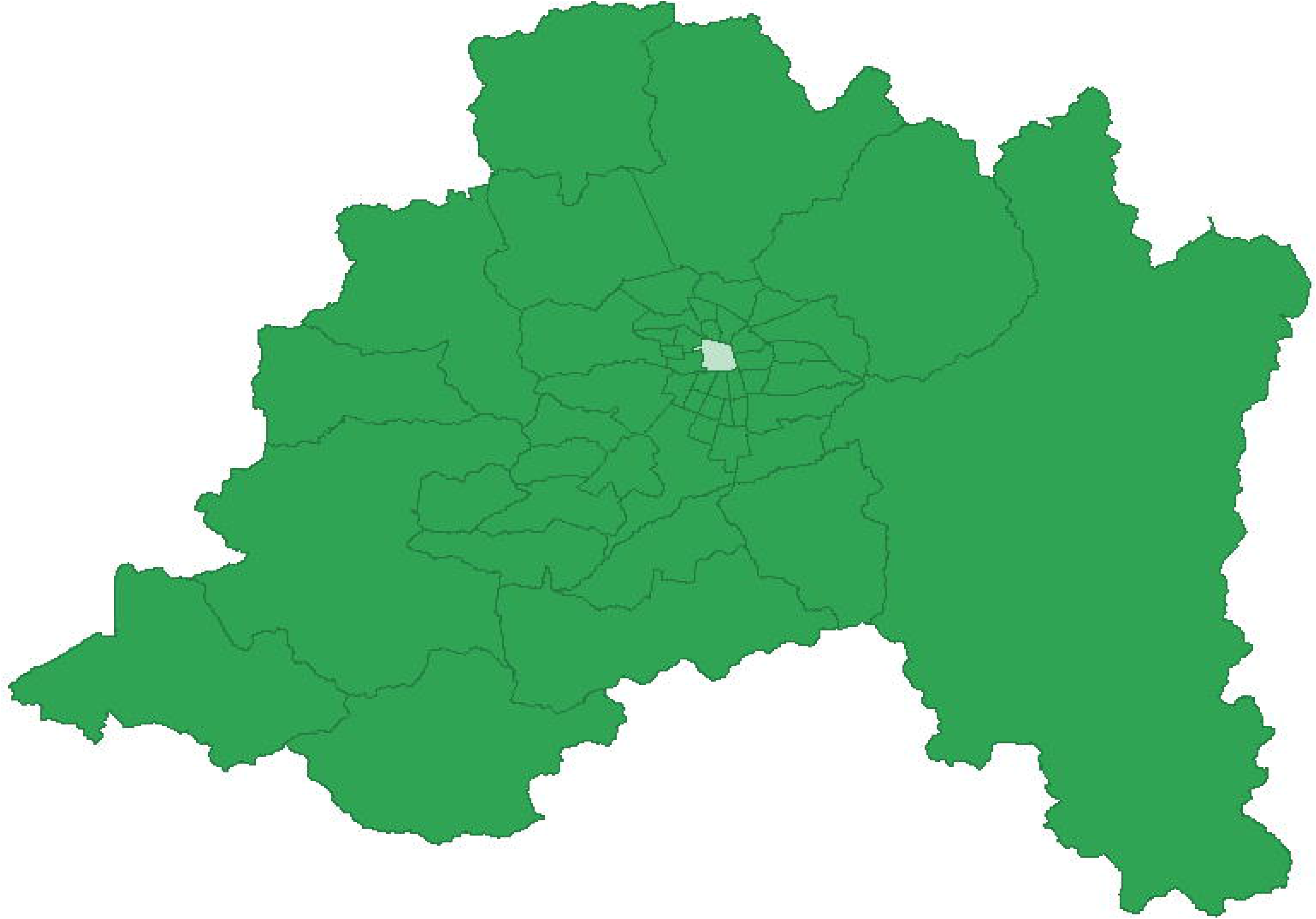
Administrative map (municipal-level) of the Metropolitan Region, Chile. Source: Authors.

On the other hand, both COVID-19 outcomes and variables of interest, particularly sociodemographic variables are not randomly distributed in the MR. People cluster based on socioeconomic and demographic characteristics, such as income and age (15-19). In Chile, several studies show marked patterns of residential segregation and geographical concentration of income, particularly in the case of the MR (19-23). There is also evidence of regional inequities in access to healthcare (24-30). Space and place have increasingly been used to analyze and understand health decisions and outcomes (31,32); this relationship has already proven to be important for the new COVID-19 disease (33-38). Understanding some of these features behind the infection and death patterns—in the context of an unequal and segregated city—is important for people, health-workers and policymakers, to address better prevention policies and community tailored intervention care programs. The information can also be relevant for other countries that, like the Metropolitan Region in Chile, are dealing with COVID-19 consequences and inequities.

## Materials and Methods

### Data sources and variables

#### Dependent variables

The study proposes the analysis of the interactions of several types of determinants and COVID-19 outcomes at the municipality level. To carry out this analysis, two groups of dependent variables were used: infections and deaths. Both variables tell a different story about the COVID-19 outcomes: while cases are mostly related to the ability to prevent infection in each municipality, deaths reflect people’s response to the virus but also how the environment (including the health system’s capacity) influences the negative consequences of the disease.

To assess the COVID-19 impact, each one of the variables was measured considering two perspectives: level and change. The two approaches allow us to look at different ways in which the disease’s impact can be measured. The level variables capture how the “length” of COVID-19 impact on the population in a particular geographical area. Change indicators provide a different measure of impact, related to the “speed” at which COVID-19 hit a determined area: not only is it relevant how many people were infected and died but also whether this happened over a short or a large period. Quick outbreaks are a sign of municipalities’ vulnerability and inability to prevent the spread of the virus but also can be relevant to understand patterns in deaths, e.g. if sudden outbreaks collapse the healthcare system. Table 1 summarizes this information.

**Table 1.** Indicators, dimensions, and dependent variables.

| Indicator/ dimension | Level | Change |
| --- | --- | --- |
| Infections | -Cumulative incidence rate (cases) after 100 days<br>-Peak of cases (daily) | Days to peak (cases) |
| Deaths | -Cumulative incidence rate (deaths) after 100 days<br>-Peak of deaths (daily) | Days to peak (deaths) |

Source: Authors.

Six different variables of interest were used in the analysis. Cumulative incidence was calculated as the number of COVID-19 confirmed cases per 100,000 people in each municipality. On the one hand, the peak variables were defined as the highest number of current (active) cases according to symptom onset date in each administrative unit. On the other hand, the speed of the impact was measured by the number of days between the first case and the maximum number of cases and deaths registered over the 100-days period for each municipality (see Supplementary Material 1). Other measures of speed, as the average growth rate in the number of cases and deaths, required a level of detail in the data (daily basis information at municipality level) that was not available.

Data on the number of positive COVID-19 cases was retrieved from open-source statistics published by the Chilean Ministry of Science, Technology, Knowledge and Innovation (39). The number of deaths was obtained from the Information and Statistic Department by the Chilean Ministry of Health (40).

#### Independent variables

To identify the effect of municipality-level features on COVID-19 outcomes, three broad groups of variables were used: demographic, health, and socioeconomic variables. The main source of information is the 2017 National Socioeconomic Characterization survey (CASEN). The CASEN survey is carried out periodically by the Chilean Ministry of Social Development since 1990, and its goal is to provide information on the socioeconomic conditions of Chilean families and assess the impact of social policies in the country. The survey includes a sample of almost 70,000 households for the 16 administrative regions in the country. This survey is based on self-reported data. For the MR, the survey contains 42,601 individual responses (41).

Demographic indicators included the share of women and older people (65+ and 80+) in each municipality. These variables have shown to be related to COVID-19 outcomes in other countries (42-46). In Chile, age is highly correlated to COVID-19-related hospitalizations and deaths (13). On the other hand, although evidence is not clear yet, the percentage of children could be relevant to explain the spread of the virus as studies have found they could have a higher viral load (47,48). They also could face COVID-19-related health complications (49,50). Additionally, other variables, such as the share of migrants, the population density, and rurality were included, as they can explain both infection rates and mortality (51-54).

A second set of variables capture health-related factors, including the health system’s features and health outcomes. As stated before, these variables have also been used in trying to identify patterns of contagion and deaths, mostly by reporting pre-existing health conditions that could relate to COVID-19 and its consequences. First, we included the population covered by the public health insurance, a self-reported variable that captures people declaring having trouble getting healthcare, and a dummy variable that identifies people that lives far from a health center (2.5 kilometers or more). These variables are expected to explain mainly COVID-19-related health outcomes (deaths), as they are proxies for healthcare access and quality and also reflect other broader inequalities in the population. A dummy variable was constructed to capture whether individuals report having at least one COVID-19-related health conditions, as reported by the Centers for Disease Control and Prevention (CDC) and the information reported by the Chilean Ministry of Health, (13, 57, 58).

Finally, several variables that capture the socioeconomic level were considered. First, water availability can be seen both as a socioeconomic and health indicator. Given the nature of the virus, access to water is expected to impact people’s preventive behavior, particularly when hand washing has been identified as a key prevention strategy for the COVID-19 since the start of the pandemic (59-60). Second, we included poverty as an indicator that captures several risk factors associated both with higher rates of infection and death. Poverty is associated with people’s vulnerability to COVID-19 and impacts the variables of interest through several channels: poor sanitary conditions, access to information, inability to follow prevention strategies—such as handwashing, the use of facemasks, and social distancing—, and lower access to healthcare, among others. (61-65) The CASEN survey includes a variable called multidimensional poverty, an index of socioeconomic vulnerability that summarizes several aspects related to social determinants and COVID-19 (see Supplementary Material 2). Using this index has advantages and disadvantages. On the one hand, it simplifies the estimation by capturing several socioeconomic factors into one variable, as used in other studies (68, 69). On the other hand, it hinders the understanding of the underlying channels through which social determinants impact health outcomes. As highlighted by Victor Fuchs “If we constantly redefine poverty to include anything and everything that contributes to poor health, we will make little progress either in theory or practice.” (66). Consequently, we estimate models including this index and others in which the different dimensions—income poverty, overcrowding, education, health insurance coverage, and job status—are considered independently. All these variables can explain infections and deaths by capturing the household’s structural inability to follow the preventive measures (handwashing, use of face masks, physical distancing, and quarantines) and seek healthcare (e.g. health literacy) (70-75). Finally, the use of public transportation and the availability of green spaces capture people’s ability to stay at home and comply with social distancing strategies. These are expected to impact the infection rate but can also influence COVID-19 deaths, since they also show difficulty to move within the city, a factor that could be relevant in case of health emergencies (76,77)

Finally, maps and spatial information were obtained from the website of the Inter-Ministerial Committee on Geographic Information (78).

All dummy variables were transformed to capture the percentage of people reporting each condition in each municipality. Continuous variables represent the municipality’s average value. The survey’s expansion factor at municipal-level was used to expand the sample’s values to a population estimate. A detailed definition of each variable is presented in Supplementary Material 3.

The selected variables cover an ample spectrum of dimensions (demographic, health, and socioeconomic factors) related to the impact (infection and deaths) of COVID-19. In terms of the spatial analysis, they include variables to explain geographical variations based on compositional issues (i.e. differences in the kind of people who live in each place) as well as contextual explanations (i.e. differences between the places), understanding that both are relevant in the relationship between health and place (79).

### Data analysis

To acknowledge the temporal dynamics of the disease, the study uses a standardized 100-days period since the first case was reported in each municipality. As for the role of space and place in explaining COVID-19 cases and deaths, we used a spatial analysis approach to look at the data.

To identify the determinants of infection and mortality due to COVID-19 in the Metropolitan Region in Chile, we carried out multivariable regressions to explain the set of dependent variables, using the three groups of explanatory variables described above. As previously stated, one of the most salient features when performing municipality-level analysis in the Metropolitan Region in Chile is the presence of geographic clusters, particularly when looking at socioeconomic indicators. Considering the way in which COVID-19 is transmitted and the potential differences in access/quality of healthcare, we expect space and place to play an important role in determining both, values in the independent and dependent variables, as well as their interactions. This justifies the use of spatial analysis in the study.

The first step in the spatial analysis is the definition of neighbors. In this case, we used a first-order queen contiguity matrix, i.e. we defined as neighbors all municipalities that share a border. Given the nature of the data—the existence of clusters in different areas of the region and the high heterogeneity in the size of the municipalities—a distance-based approach was discarded (79).

To understand the impact of different variables in the incidence and mortality due to COVID-19 in the region, we use ordinary least squares (OLS) regressions. If space and place are relevant in determining the variables in our analysis, then the errors in the OLS regressions are spatially-correlated and, consequently, the results are biased (80). To test the presence of spatial correlation on the OLS regression, the global Moran’s I test—that indicates both the existence and degree of spatial autocorrelation (82)— is applied to the regression’s residuals. This test shows whether the residuals are randomly distributed. If the null hypothesis is rejected, the OLS analysis needs to be adjusted to consider the spatial effects. There are two main strategies to estimate spatial autoregressive models: spatial lag and spatial error. The spatial lag model (also known as contagion model) incorporates space as a right hand-side variable, estimating a coefficient for the spatial effect; the error model does not incorporate spatial as a covariate, but includes it in the structure of the residuals (81). Conceptually, spatial lag models seem more appropriate to adjust for spatial autocorrelation in the case of infections, since it is expected that the number of COVID-19 cases in one municipality affects the cases in the neighboring areas, the contagion effects. However, this is not necessarily true for mortality, particularly once controlling for the case incidence rate. In this case, a spatial error model appears more suitable, since unobserved spatial effects are expected to drive the spatial autocorrelation in the residuals. Consequently, spatially correlated regressions of cases are adjusted using a spatial lag model, and an error model is utilized for the death regressions.

Incidence and mortality data were collected and calculated using Microsoft Excel. Descriptive statistics and multivariable regressions were estimated using STATA, and spatial analysis and visualizations were conducted in GeoDa.

## Results

Table 2 shows the descriptive statistics of the sample. First, it is observed the large heterogeneity in most variables between municipalities, both in the dependent and independent variables. As stated before, this reflects the different realities within the MR, as well as the differences in terms of COVID-19 outcomes. Second, spatial autocorrelation appears as statistically significant for all the variables of interest (different measures of infection and mortality), as well as many of the independent variables, particularly those related to social determinants of health.

**Table 2.** Descriptive statistics,

| Variable | Average | St. Dev. | Min | Max | Moran's I |
| --- | --- | --- | --- | --- | --- |
| Cumulative incidence rate (cases) | 2,477.00 | 901.02 | 1025.10 | 4,788.00 | 0.43*** |
| Peak of cases | 2,106.99 | 857.47 | 588.00 | 3,983.90 | 0.43*** |
| Days to peak of cases | 93.00 | 11.53 | 65.00 | 118.00 | 0.16** |
| Cumulative incidence rate (deaths) | 92.55 | 45.39 | 25.10 | 224.25 | 0.46*** |
| Peak of deaths | 50.79 | 29.74 | 3.51 | 137.85 | 0.18** |
| Days to peak of deaths | 84.27 | 15.47 | 38.00 | 107.00 | 0.10* |
| Women | 0.52 | 0.02 | 0.46 | 0.56 | 0.04 |
| Children | 0.07 | 0.01 | 0.04 | 0.09 | -0.04 |
| People 65+ | 0.13 | 0.04 | 0.06 | 0.23 | 0.26*** |
| People 80+ | 0.03 | 0.01 | 0.00 | 0.07 | 0.07 |
| Migrants | 0.03 | 0.06 | 0.00 | 0.38 | 0.26*** |
| Population density | 6,165.66 | 6,152.61 | 3.73 | 22,870.32 | 0.65*** |
| Rurality | 0.12 | 0.21 | 0.00 | 1.00 | 0.48*** |
| Public health insurance | 0.75 | 0.18 | 0.13 | 0.99 | 0.49*** |
| Problems accessing healthcare | 0.00 | 0.00 | 0.00 | 0.01 | 0.03 |
| Distance to health center | 0.88 | 0.13 | 0.45 | 0.99 | 0.29*** |
| COVID-19 risk factors | 0.16 | 0.03 | 0.07 | 0.24 | 0.24*** |
| Water inside the house | 0.98 | 0.02 | 0.92 | 1.00 | 0.02 |
| Multidimensional poverty | 0.22 | 0.08 | 0.03 | 0.38 | 0.30*** |
| Income poverty | 0.06 | 0.03 | 0.00 | 0.14 | 0.26*** |
| No overcrowding | 0.89 | 0.05 | 0.78 | 0.98 | 0.27*** |
| Critical overcrowding | 0.01 | 0.01 | 0.00 | 0.05 | 0.12* |
| Years of education | 5.88 | 4.14 | 1.55 | 18.67 | 0.47*** |
| Self-employed worker | 0.10 | 0.02 | 0.05 | 0.15 | 0.06 |
| Green spaces | 9.63 | 1.50 | 8.00 | 14.05 | 0.33*** |
| Use of public transportation | 0.21 | 0.07 | 0.05 | 0.41 | 0.49*** |

Note: All variables are expressed as the share of the municipality's total population, except for "Population density", "Green spaces", "Years of education" and "Minutes in public transportation" that report the municipality's average. Min and Max refer to the minimum and maximum values at municipality, not individual level. Significance level: \*\*\* $p < 0.01$ , \*\* $p < 0.05$ , \* $p < 0.1$ . Moran's I statistical significance was assessed using pseudo p values with 999,999 permutations.

This result gives a first warning on the potential effect of space between COVID-19 outcomes and municipal-level features. Spatial autocorrelation and measures can be broadly classified into global and local measures (83). Moran’s I is a global test that indicates the presence of spatial correlation; a local test can be used to answer where this correlation is. Fig 2 shows the results for the Gi* tests in selected dependent and independent variables. The Gi* identify areas where the values of the variables are high or low (hot and cold spots detection) respect to the global average (83). The figure exhibits clusters around the Santiago downtown area vs peripheric municipalities, as well as an east-west pattern, particularly for socioeconomic variables.

**Fig 2.**
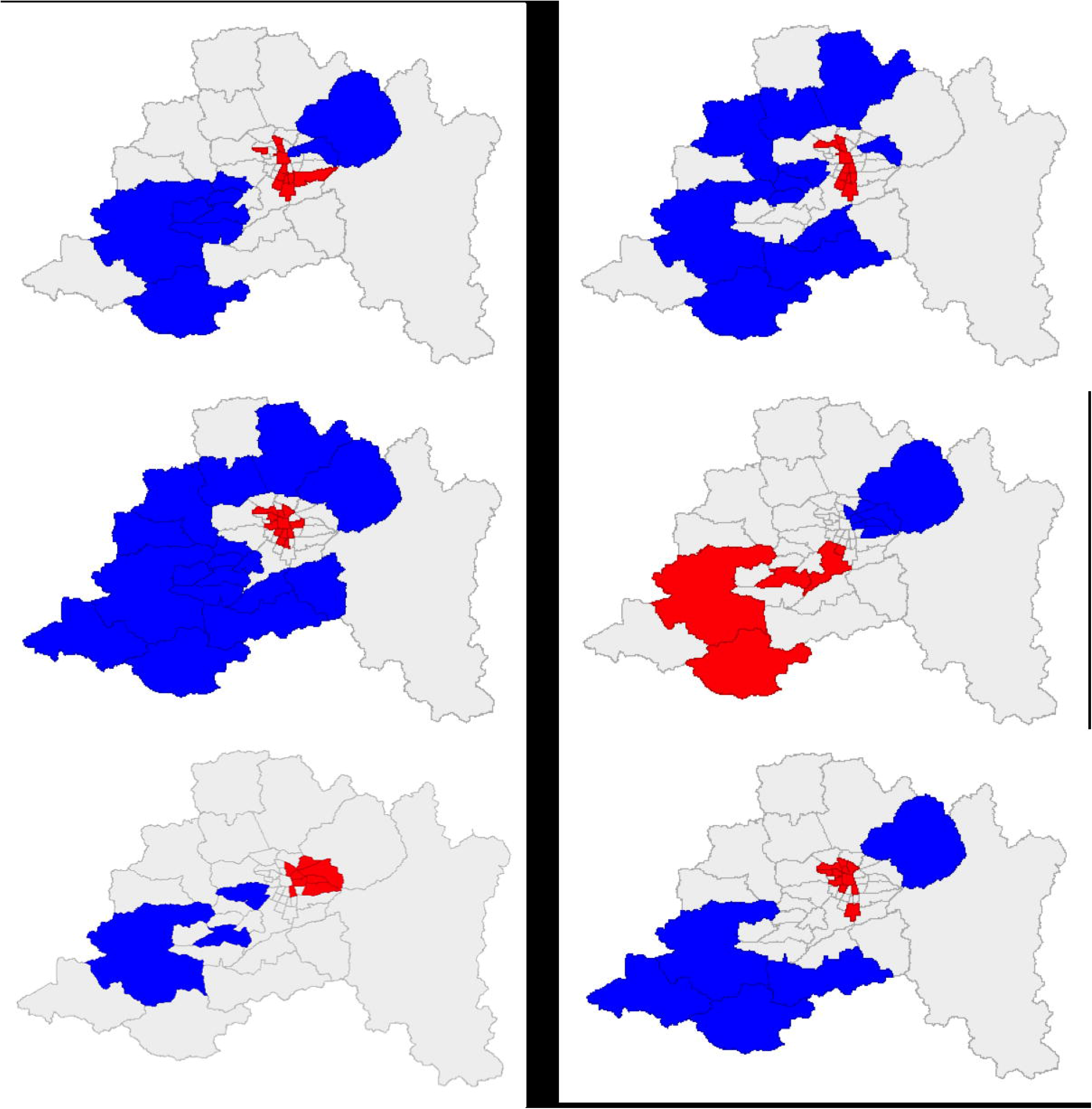
Gi* tests for clustering in selected variables Note: Upper-left: infections (100 days cumulative rate); Upper-right: deaths (100 days cumulative rate); Middle-left: population density; Middle-right: public health insurance; Down-left: average years of education; Down-right: use of public transportation. Blue: low-value clusters (95% significance); Red: high-value clusters (95% significance); Grey: not significant areas.

To identify how different variables relate to COVID-19 infections and deaths, several multivariable regressions were estimated. Table 3 presents the results for the infection-related dependent variables. As shown in Table 1, each variable has three different ways to be measured: cumulative incidence rate (columns 1 and 2), peak of cases (columns 3 and 4), and days to the peak (columns 5 and 6). Each model is estimated using multidimensional poverty (even columns) and a set of socioeconomic variables (odd columns).

**Table 3.** Infection regressions (OLS)

| VARIABLES | Cumulative incidence rate |  | Peak |  | Days to peak |  |
| --- | --- | --- | --- | --- | --- | --- |
|  | (1) | (2) | (3) | (4) | (5) | (6) |
| Women | -5.434 | -6.097 | 4.462 | 4.158 | 0.115 | 0.144 |
| Children | -0.043 | -3.667 | 6.027 | 0.792 | 0.0805 | 0.073 |
| People 65+ | 6.415 | 5.517 | -2.984 | -4.732 | **-0.154\*\*** | **-0.135\*** |
| People 80+ | 0.488 | 6.113 | -1.859 | 1.479 | -0.0211 | -0.000521 |
| Migrants | -2.751 | -5.453 | 1.504 | 1.268 | 0.0231 | 0.0429 |
| Population density | 4.07E-05 | **6.35e-05\*** | 2.45E-05 | 4.59E-05 | -3.33E-07 | -4.92E-07 |
| Rurality | 0.89 | 1.547 | -0.176 | -0.267 | -0.025 | **-0.044\*\*\*** |
| Multidimensional poverty | **5.065\*\*\*** |  | **4.528\*\*\*** |  | 1.68E-05 |  |
| Income poverty |  | 2.433 |  | 5.885 |  | -0.0148 |
| No overcrowding |  | -2.11 |  | 2.701 |  | -0.00375 |
| Critical overcrowding |  | 10.98 |  | 8.536 |  | -0.0337 |
| Years of education |  | -0.101 |  | -0.33 |  | 0.00116 |
| Self-employed worker |  | 4.467 |  | 0.117 |  | **-0.198\*\*** |
| Green spaces | 0.0232 | 0.0223 | -0.0161 | -0.00561 | **-0.000915\*** | -0.00069 |
| Use public transportation | **9.526\*\*\*** | **8.803\*\*\*** | 2.537 | 2.122 | -0.0459 | -0.0606 |
| Water inside the house | -4.636 | -2.112 | -0.262 | -1.411 | 0.0485 | 0.0593 |
| Public health insurance |  | 0.893 |  | -0.86 |  | 0.0164 |
| Difficulty getting healthcare | -91.34 | **-113.2\*** | 30.93 | 43.17 | 1.279* | **1.957\*\*** |
| Distance to health center | 0.441 | 0.379 | 1.347 | 1.194 | 0.00237 | -0.0095 |
| COVID-19 risk factors | 0.0994 | -3.177 | 4.436 | 3.435 | 0.028 | 0.0604 |
| Constant | 5,196 | 6,448 | -3,440 | 490.1 | 12.54 | -5.576 |
| R-squared | 0.665 | 0.692 | 0.609 | 0.632 | 0.351 | 0.428 |
| Adjusted R-squared | 0.538 | 0.509 | 0.461 | 0.414 | 0.105 | 0.088 |
| Moran's I (residuals) | -0.0374 | -0.0222 | -0.0587 | -0.0687 | -0.0309 | -0.0196 |

Significance level (robust standard errors): \*\*\* p<0.01, \*\* p<0.05, \* p<0.1

First, it is noted that the determinants of level and change in infections differ. As expected, the use of public transportation shows a significant and positive association with cumulative incidence rate; results for change-type regression (columns 5 and 6) are less consistent, with the share of people 65+, rurality, self-employment, green spaces, and difficulty to get healthcare showing significant coefficients. Second, multidimensional poverty appears as important to explain the number of cases (columns 1 and 3), while the effect vanishes when looking at the impact of a set of socioeconomic variables instead of the index of socioeconomic vulnerability (columns 2 and 4). The effect, as expected, is also positive showing that multidimensional poverty is a risk factor for COVID-19 contagion at the municipality level. Third, the model’s overall fit is better in the leveltype regressions, explaining 60% of the variation in the dependent variables. In the case of estimations 5 and 6, count models using Poisson regression were also estimated; results—in terms of significance and sign of the coefficients—hold when using this alternative.

Finally, Moran’s I tests for spatial autocorrelation of the residuals show that the hypothesis that OLS residuals are distributed randomly in the space cannot be rejected for all models. Consequently, there is no statistical justification for adjusting to consider spatial effects.

Table 4 presents the same set of estimations for the mortality variables. In this case, four different models are estimated for each one of the independent variables, cumulative mortality rate (columns 1 to 4), peak of deaths (columns 5 to 8), and days to the peak of deaths (columns 9 to 12). As before, each model is estimated using either the multidimensional poverty index or a set of socioeconomic factors; additionally models are estimated including and excluding the cumulative incidence rate of cases as explanatory variable (even and odd columns, respectively).

**Table 4.**
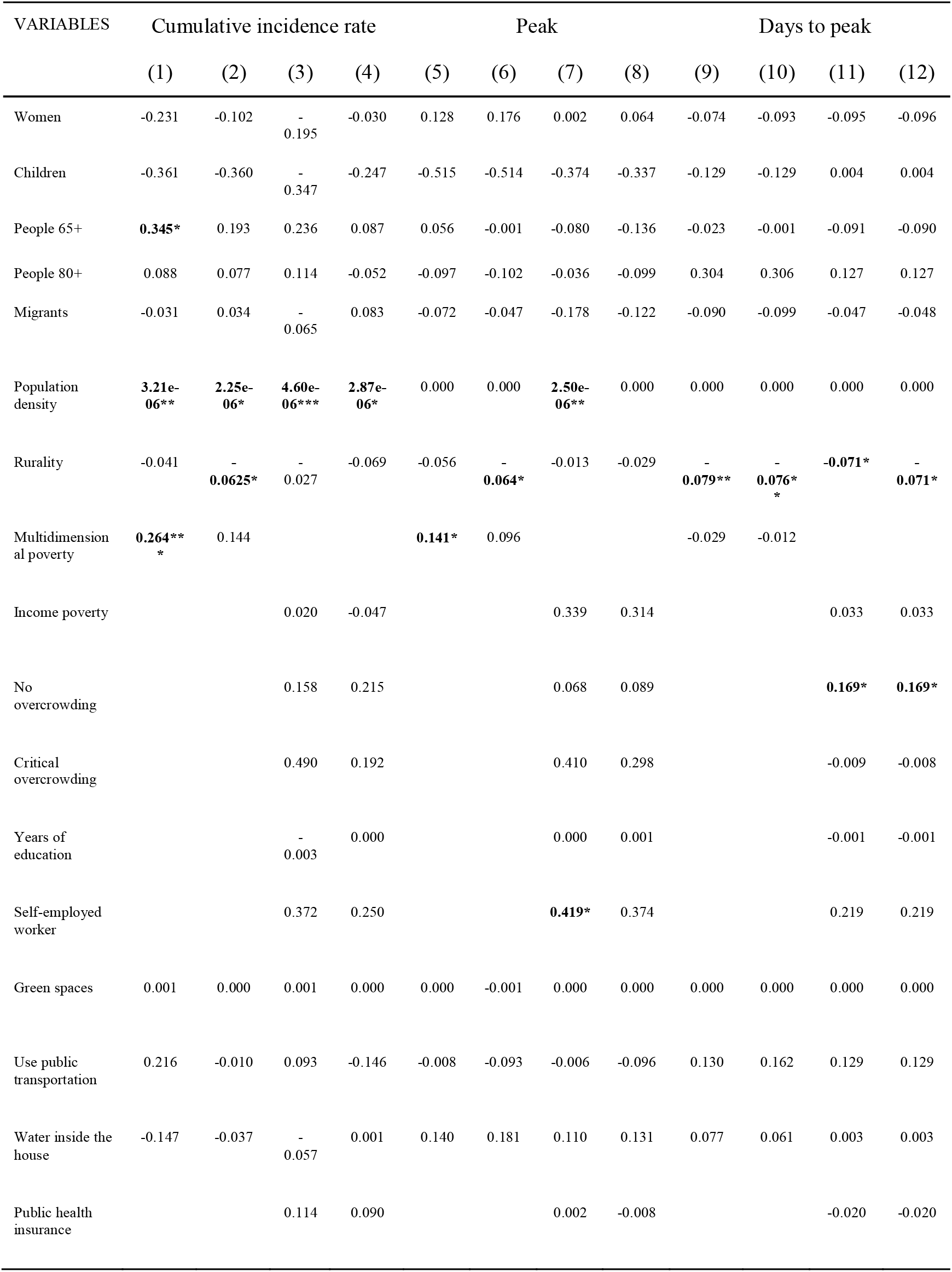
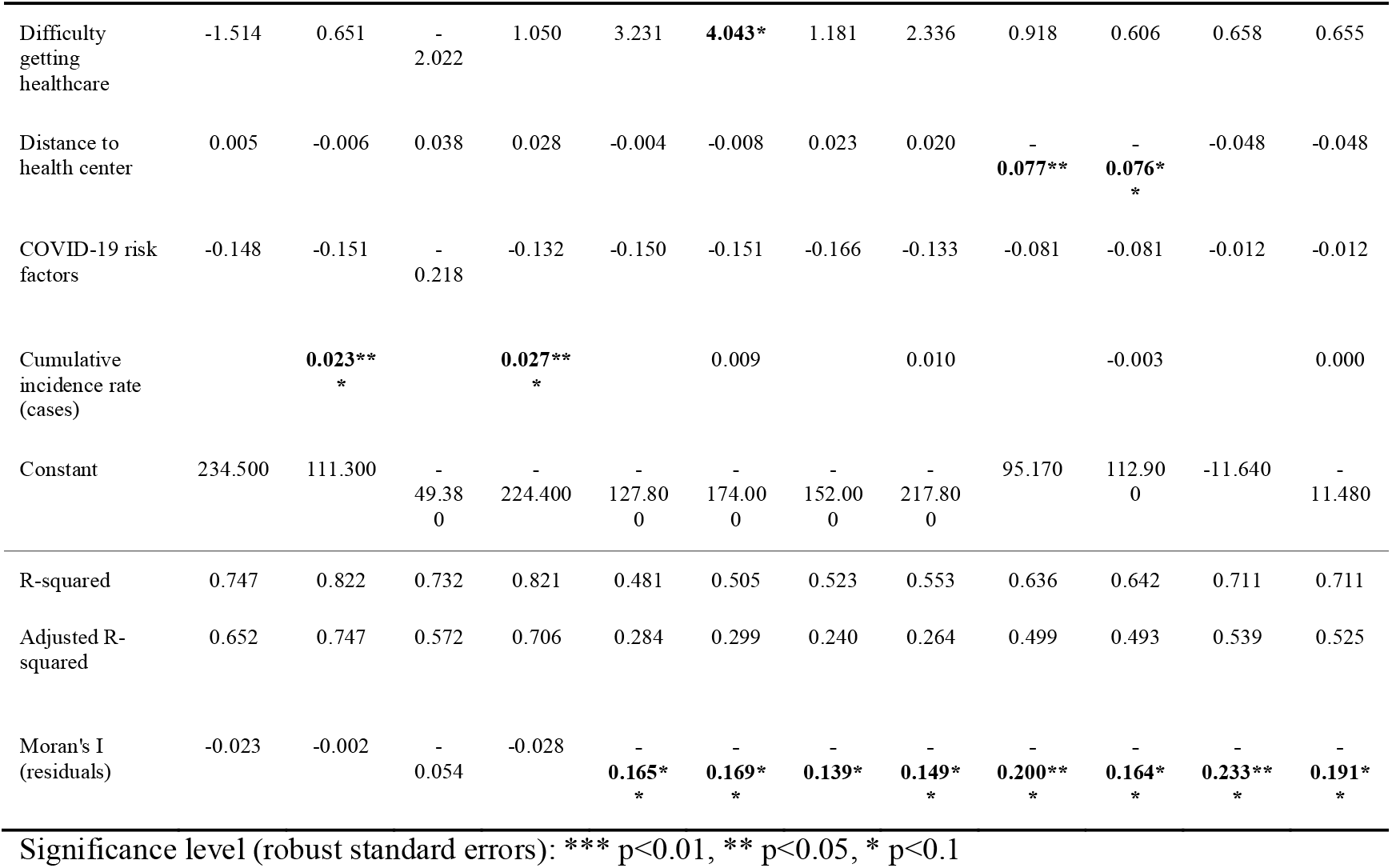
Deaths regressions (OLS)

The first result is that the determinants of infections and deaths are not the same. However, as expected, some variables seem to explain variation in both types of variables. As in the case of infections, level-type and change-type estimations present common and specific determinants.

For deaths, for level-type variables (columns 1 to 8) the share of people over 65 years old, population density, multidimensional poverty, and the prevalence of cases have significant and positive coefficients; overcrowding and distance to a health center also contribute to explain whether a municipality reaches the peak of cases faster or slower. Just like in the case of infection models, multidimensional poverty captures an effect that is not explained by a broad set of socioeconomic factors. The model also does a better job explaining cumulative rates than peaks and days to the peak, and the overall fit (R^2^) is larger than for the infection regressions in Table 3. As before, using Poisson regressions for days to the peak does not change the main results.

Unlike the infection regressions, in this case, the hypothesis of a random spatial distribution of the residuals is rejected in 8 out of 12 cases, highlighting the need to use spatial regression models to take into account the presence of spatial autocorrelation.

Based on the results from Tables 3 and 4, a simplified set of regressions is estimated. A potential problem with inference, in this case, is related to the degrees of freedom due to a large number of explanatory variables and the relatively small sample (n=52). Table 5 shows these reduced models, based on the previous results (infection and deaths OLS regressions). As emphasized above, both variables (cases and deaths) share some explanatory variables and differ in others. In this case, it is also observed that level-type and change-type models have different determinants. Notably, the overall fit of the model using a reduced set of independent variables is similar to the one obtained in Tables 3 and 4, showing that an important proportion of the variation in the variables of interest can be explained by a small set of variables.

**Table 5.** Simplified regressions (OLS)

| VARIABLES | Cumulative<br>infection<br>rate | Peak of<br>cases | Days to<br>peak (cases) | Cumulative<br>death rate | Peak of<br>deaths | Days to peak<br>(deaths) |
| --- | --- | --- | --- | --- | --- | --- |
|  | (1) | (2) | (3) | (4) | (5) | (6) |
| People 65+ |  |  |  | **0.147\*** | 0.0923 | 0.027 |
| Population density | **4.25e-05\*** | **3.19e-05\*** | -3.82E-07 | **2.61e-06\*\*\*** | 8.96E-07 | 2.12E-07 |
| Rurality |  |  |  | **-0.0568\*\*\*** | **-0.070\*\*\*** | **-0.114\*\*\*** |
| Multidimensional<br>poverty | **5.691\*\*\*** | **5.185\*\*\*** | 0.00244 | 0.115* | 0.0905 | -0.0212 |
| Use public<br>transportation | **4.697\*\*\*** | **5.076\*\*\*** | **0.0688\*\*** |  |  |  |
| Difficulty getting<br>healthcare | -31.23 | 4.56 | -0.175 |  |  |  |
| Distance to health<br>center |  |  |  | -0.00164 | -0.0229 | **-0.0943\*\*\*** |
| Cumulative<br>incidence |  |  |  | **0.0225\*\*\*** | 0.00523 | -0.000648 |
| Constant | 33.57 | -280.9 | **80.76\*\*\*** | -15.59 | 29.01 | **155.0\*\*\*** |
| R-squared | 0.568 | 0.562 | 0.101 | 0.805 | 0.423 | 0.576 |
| Adjusted R-squared | 0.532 | 0.525 | 0.025 | 0.780 | 0.346 | 0.519 |
| Moran's I (residuals) | 0.081 | -0.087 | **0.1251\*\*** | -0.0056 | -0.1118 | **-0.1418\*** |

Significance level (robust standard errors): \*\*\* p<0.01, \*\* p<0.05, \* p<0.1

Finally, considering the presence of spatial autocorrelation in the OLS residuals, spatial regressions are used to control for these effects. As discussed above, a spatial lag regression is used for the infection model (column 1), while spatial error regressions are estimated for death models (columns 2 to 10). In both cases, the same specification used in Tables 2 to 5 was utilized.

First, adding a spatial dimension removes the spatial correlation in the residuals in six cases where OLS residuals show spatial autocorrelation: days to peak of cases (column 1), peak of deaths (columns 2 to 5), and days to peaks of deaths (columns 6 to 10). However, overall the results improve, reflecting the addition of a previously omitted significant variable. Not only overall fit increases (R^2^) but also results in terms of individual coefficients become more consistent. As Table 6 shows, the percentage of children and the rurality reduces the magnitude in the peak of deaths but makes it quickly (reduce the days to the peak). The opposite occurs for multidimensional poverty, again, a risk factor to explain the level and velocity of deaths. The speeding effect is also observed for the percentage of people 65+, migrants, overcrowding, and distance to health centers, while the years of education increase the number of days to reach the peak. As before, population density shows positive and significant coefficients for the peak of deaths regressions.

**Table 6.**
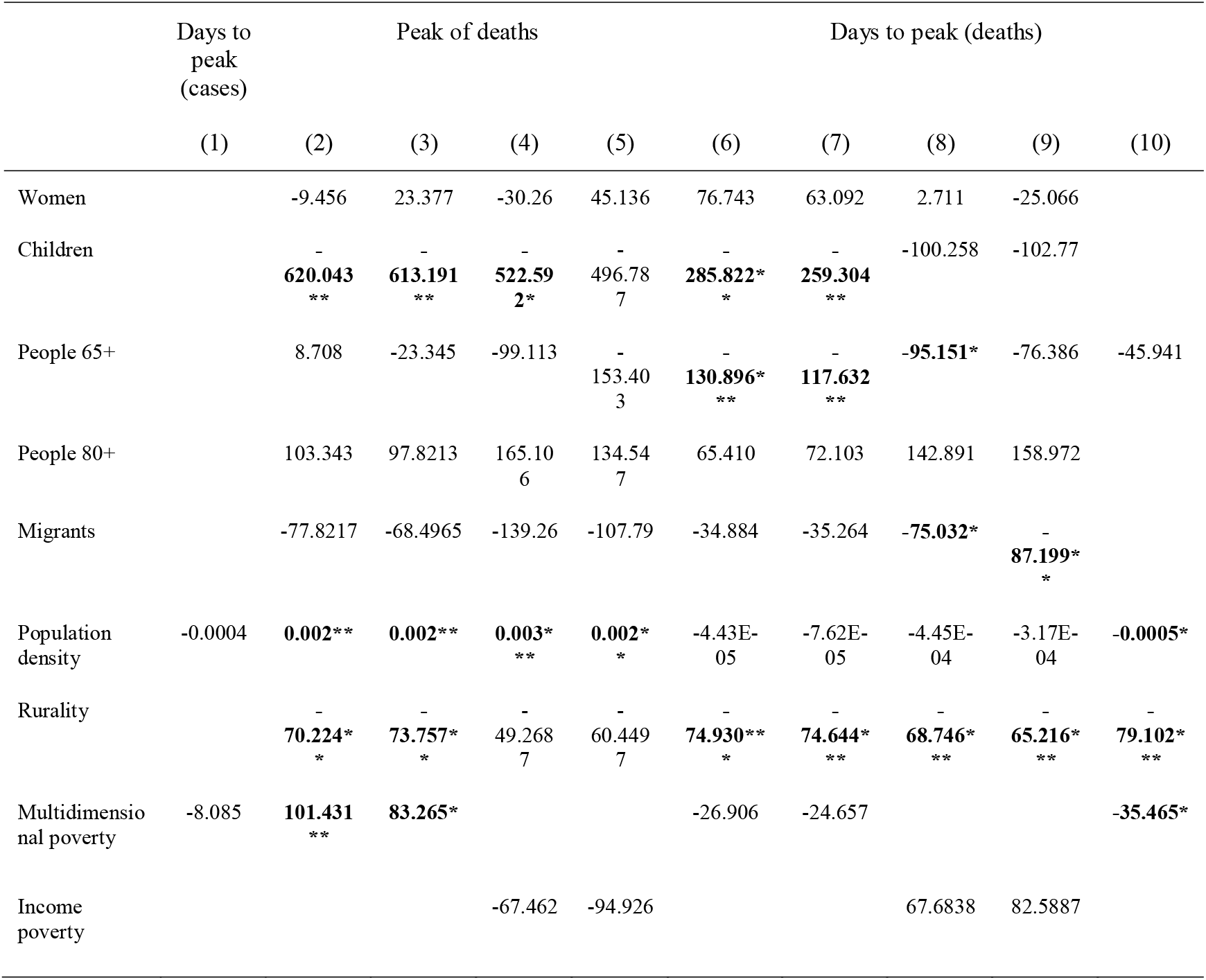
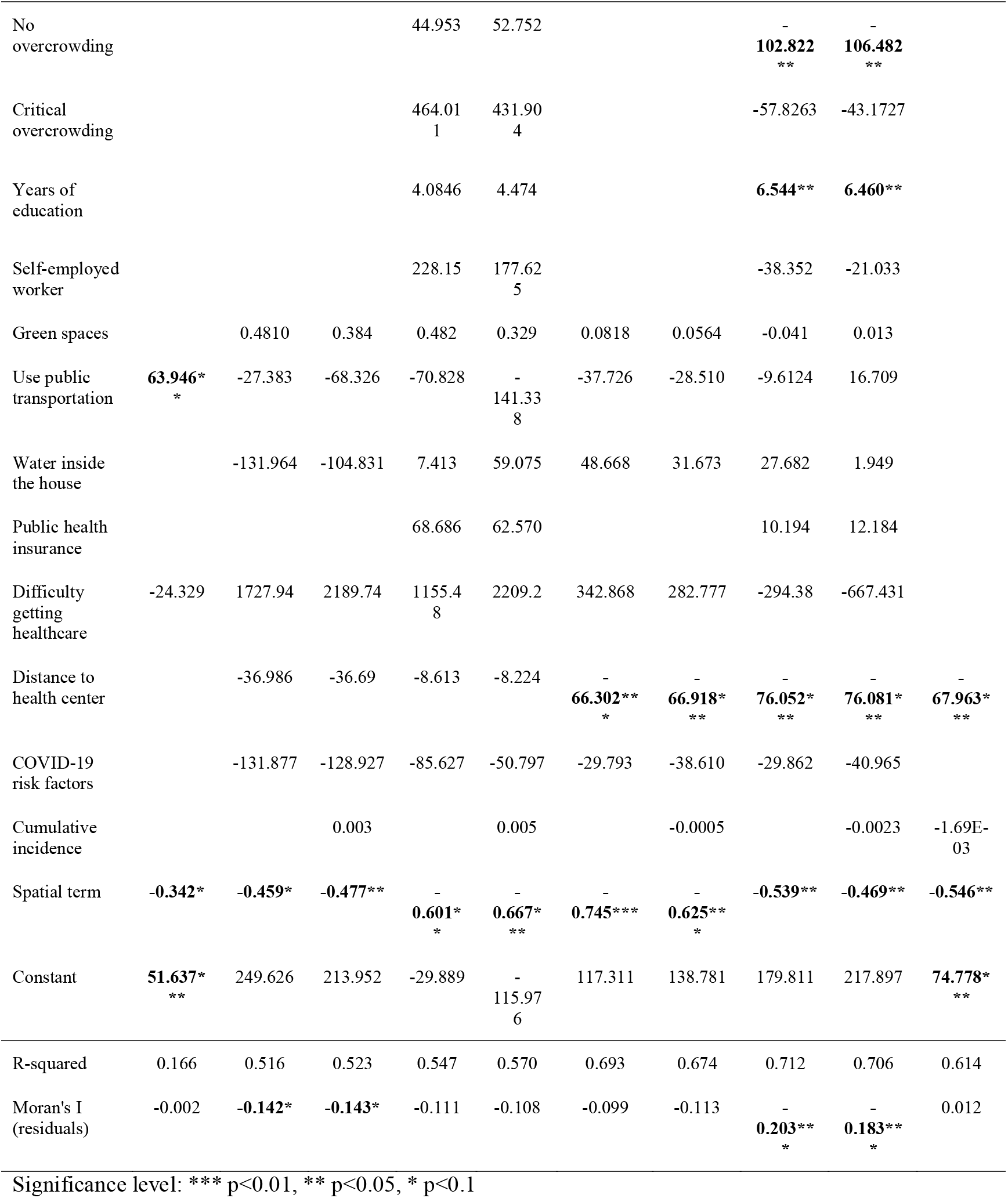
Spatial regressions

## Discussion

Using municipality-level data on 52 administrative units, the article explored the effects of different sets of variables, acknowledging the need to consider a broad set of variables, as well as time dynamics and spatial effects in the analysis.

Several conclusions are drawn from the results. First, there are common and idiosyncratic elements that explain the prevalence and dynamics of infections and mortality. It is necessary to recognize these different approaches when discussing the “impact” on COVID-19. In this line, the proposed models are better explaining variation in levels of infections and deaths than changes (measured as days to the peak). Research on the outcomes of COVID-19 needs to incorporate different perspectives (measures of impact and determinants) to shed light on the problem. Second, as long as different models are required to tell different stories (34 estimations in this case), a significant part of the municipal variation in infections and deaths can be explained by a rather small number of variables (as in Table 5). In any case, all the results highlight the role of social determinants of health in explaining the dissimilar impact of COVID-19 in the MR as well as other findings in other countries about unequal distribution of COVID-19 (83-86).

In this line, the use of multidimensional poverty index as a determinant of COVID-19 outcomes is an interesting result; the indicator shows to be relevant to explain infections and deaths, doing a good job capturing the complex nature of the problem, highlighting the role played by structural determinants—particularly with poverty and vulnerability— and showing similar outcomes to other studies (87-90)This result has more relevance considering that COVID-19 also has an impact on socioeconomic factors, generating a vicious circle between both problems (91-93). Third, the results identify different types of variables that explain the COVID-19 outcomes: demographic, health-related, and socioeconomic. However, these determinants can also be grouped according to their degree of changeability. Some of the relevant indicators can be seen—at least in the short run—as fixed (such as poverty and age distribution), while others are more likely to be changed (like the use of public transportation and the availability of a health center). From a public policy perspective, this classification is useful: the first group of indicators can be assumed in the short-term as “explanatory” but be used for long-term planning; on the other hand, the second group of indicators are “policy tools” and can be used as control knobs to short-term responses to the pandemic.

The article has some limitations that need to be taken into account when interpreting the results. First, there are time-differences between dependent and independent variables: while dependent variables reflect information between March and July 2020, independent variables come mainly from a survey carried out three years before. Unfortunately, there is no other municipality-level representative source with more updated information. However, most of the variables are expected to be similar today. There are at least two reasons for this. First, aggregated data is expected to change slower than individual-level data. Second, as discussed above, most variables reflect structural factors—such as demographic and socioeconomic features—that are not expected to change significantly in a three-year period. This is also expected for “policy tools” variables (such as the patterns of use of public transportation) in absence of a policy shock. Second, there is a difficulty in choosing the concrete way to measure the relevant outcomes. Of course, different specifications can lead to different results. In this analysis, the issue was confronted by using several perspectives (infections and deaths, and levels and change). In a similar vein, methodological choices are also required when selecting the unit of analysis. In this case, we focused on municipalities and used percentages of individuals (instead of, for example, households) to calculate the independent variables. Finally, regarding spatial analysis, the definition of neighbors is also a crucial one; although in this case the selection of contiguous units is justified by the features of the data (the existence of units of different sizes and the spatial patterns of the variables of interest). The Metropolitan Region is a good case for spatial analysis but other geographical areas could also be of interest to understand the COVID-19 dynamics and policy responses (e.g. for establishing regional or international sanitary customs).

## Conclusions

This article presents a multi-perspective analysis of the COVID-19 impact in the Metropolitan Region, Chile. It provides several interesting insights for understanding the patterns and dynamics of the disease, and highlights the need to look at social determinants of health and spatiotemporal dynamics in the analysis of COVID-19 (94). It also opens a large space for further research questions that could not be answered here. One important question related to the outcomes and relationships once the pandemic is finished. The use of a 100-days period allows us to understand the evolution of the disease so far but it is a snapshot of an ongoing issue. Additionally, better availability and quality of data can help to understand the underlying factors behind the changes in COVID-19 outcomes. We proposed an imperfect indicator for measuring the speed of changes. There are still relevant questions to answer in this line. Finally, the estimations are based on municipalities’ structural features; the understanding of COVID-19 outcomes can also improve by adding real-time variables related to people’s behavior, such as the percentage of people using face masks, or better data on mobility within and between geographical areas.

We believe the analysis contributes to the worldwide effort for understanding the social determinants and effects of the COVID-19. Our results can be used to promote health in all policies and to enhance intersector efforts to face this pandemic. We expect these results to be particularly useful for policymakers, and hope they can encourage the implementation of evidence-based solutions and tailored interventions to help minimize the negative effects of the pandemic tackling social inequalities.

## Data Availability

Data available upon request

## Acknowledgments

The authors thanks comments and suggestions from Carla Castillo-Laborde, Italo Lopez, and Felipe Elorrieta.

**Supplementary material 1**. Date used to establish the period of analysis in each municipality

**Supplementary material 2**. Index of multidimensional poverty

**Supplementary material 3**. Definition of variables

